# A Clinical study to evaluate efficacy of CaFi (Branded ingredient of Cassia Fistula Extract) in the healthy individuals having Irregular bowel habits-An Open Label, Randomized, Comparative, Multi centric, Interventional, Prospective, Clinical Study

**DOI:** 10.1101/2025.05.28.25328477

**Authors:** Sanjay Tamoli, Mrudul Chitrakar, Nilesh Londhe, Santosh Irayya Swami, Mahesh Kumar Harit, Sarika Londhe, Channamma Hiremath, Ruby Dubey, Ninad N. Mulye

## Abstract

**Background:** Irregular bowel habits including constipation are common gastrointestinal issues, significantly impacting quality of life. While conventional pharmacological treatments exist, they often come with side effects and offer limited long-term relief, driving interest towards safer, effective herbal alternatives. *Cassia fistula (C. fistula)* has long been recognized for its laxative properties and has been reported to be a safer alternative to Senna *(Cassia angustifolia)*.

**Purpose:** This study aimed to evaluate the efficacy and safety of a standardized extract of *C. fistula* compared to Senna extract in healthy individuals experiencing irregular bowel habits.

**Methods:** A two-arm, open-label, randomized, multicentric, interventional, prospective clinical study was conducted in India with 70 participants (35 per group). One group received the standardized *C. fistula* extract and the other group received Senna extract for 14 days, followed by a 7-day post-consumption follow-up. Primary outcomes included changes in bowel movement frequency. Secondary outcomes assessed stool consistency (Bristol Stool Form Scale), straining, bowel satisfaction after defecation, anorectal blockage, use of manual manoeuvres, average time spent on defecation, time to first bowel evacuation post intervention and associated clinical symptoms (headache, belching, flatulence, abdominal distension, acidity). Safety was evaluated through vitals, laboratory parameters, and adverse events.

**Results:** *C. fistula* extract significantly increased bowel movement frequency (3.30 ± 0.92 to 5.90 ± 1.24 at Day 14, p<0.05) and improved stool consistency comparable to Senna extract. Straining and bowel satisfaction after defecation also significantly improved with the use of C. Fistula. Notably, *C. fistula* extract showed significantly greater reduction in the sensation of anorectal blockage at Days 14 and 21 (p<0.05) and a faster onset of action (shorter time to first defecation) compared to Senna. Associated symptoms of constipation also reduced significantly. *C. fistula* extract was better well-tolerated, with fewer product-related adverse events compared to Senna.

**Conclusion:** *C. fistula* extract was found to be effective in irregular bowel habits, offering comparable efficacy to Senna while proving to be superior in reducing the sensation of anorectal blockage and achieving a faster time to first bowel evacuation. It also demonstrated a more favorable safety profile with significantly fewer product-related adverse events. Therefore, *C. fistula* extract represents a promising non-habit-forming herbal alternative for bowel regulation.

## Introduction

Constipation and irregular bowel habits are prevalent gastrointestinal conditions affecting individuals across all age groups, genders, and socioeconomic backgrounds. ^1,2^ Constipation is commonly characterized by the presence of one or more of the following symptoms: hard stools, infrequent bowel movements (≤ three per week), a sensation of incomplete evacuation, excessive straining during defecation, or spending excessive time on the toilet with unsuccessful attempts to pass stool.^3^ It is a widespread digestive disorder influenced by various metabolic, muscular, psychogenic and iatrogenic factors, ^4^ and is observed in around 15% of the population.^5^

Constipation adversely affects health and quality of life; however, it is often underreported and inadequately managed.^6^ Treatment is typically empirical, starting with lifestyle measures such as fiber intake, hydration and physical activity.^3^ When these fail, pharmacological agents may be used, but they often cause side effects and provide limited short-term relief. This has led to growing interest in safer, more effective alternatives like herbal therapies.^7^

*Cassia fistula (C. fistula)* is increasingly recognized as an effective laxative for managing constipation. Its fruit—a long pod containing seeds surrounded by sweet, dark pulp—is primarily used in the management of constipation due to its anthraquinone and high mucilage content. ^8,9^ These compounds are metabolized by gut bacteria into active agents that stimulate intestinal peristalsis and increase water and electrolyte secretion, softening stool and easing bowel movements. ^10^ Clinical studies have confirmed the laxative properties of *C. fistula* fruit pulp, often comparing it to conventional treatments.^11^

The present study evaluates the effect of a standardized extract of *C. fistula* in constipation and irregular bowel habits in healthy participants over a period of 14 days of use as compared to a group administered with Senna *(Cassia angustifolia)* extract.

## Materials and methods

### Study design and sites

The present study was a two arm, open label, randomized, multi centric, interventional, prospective, clinical study conducted at three sites in India:

1. D. Y. Patil University, School of Ayurveda, Department of Swasthavritta and Yoga OPD No. 06, 1st floor, Nerul, Navi Mumbai, Mumbai 400706, Maharashtra, India.
2. Nirvikar Hospital, Department of General OPD, OPD No. 1, First Floor, B Wing, Jay Ganesh Samrajya, Sector No. 3, Bhosari, Pimpri Chinchwad, Pune 411039, Maharashtra, India.
3. Department of Kayachikitsa, Seth Govindji Raoji Ayurved Mahavidyalaya, Solapur-413001, Maharashtra, India.

### Ethical considerations and informed consent

Study was conducted after approval from the Institutional Ethical Committees of the respective study sites –

- D. Y. Patil University, School of Ayurveda – DYPUSA/24/351
- Nirvikar Hospital – ECR/1876/INST/MH/2023
- Seth Govindji Raoji Ayurved Mahavidyalaya – IEC/LTR/SGR/12/2024-25

CTRI registration was done with registration number – CTRI/2024/08/072838. Consenting participants meeting the inclusion criteria were considered for the study. The study was carried out and reported adhering to CONSORT Statement.

### Details of intervention

The proposed intervention is a standardized herbal extract of *C. fistula* given in a dose of 250mg in tablet form.

### Study dosage, duration and visits

The study involved two groups; one group was administered one tablet (containing 250 mg) of standardized *C. fistula* extract at bedtime with water for 14 days. The other group (control group) was given Senna extract in tablet form at bedtime with water for 14 days.

Interventions were administered for a total duration of 14 days, followed by a 7-day post-treatment follow-up period. Study visits were planned and various outcomes were assessed on screening visit, baseline visit (day 0) and days 7, 14, 21.

### Inclusion criteria

Healthy participants of both genders aged 18 to 70 years (inclusive), presenting with two or more of the Rome IV diagnostic criteria for functional constipation, (2016)^12^ for the past 3 months, with symptom onset at least 6 months prior to screening, and who were willing to provide written informed consent and comply with protocol requirements of the clinical study were included.

### Exclusion criteria

Participants were excluded if they had colonic inertia, a history of abdominal or anorectal surgery, or other functional gastrointestinal disorders (e.g., IBS, belching disorders). Structural anorectal abnormalities, uncontrolled diabetes or hypertension, metabolic, infectious, or neurological disorders, long-term medication use (including constipation-inducing drugs), pregnancy or lactation, known hypersensitivity to study ingredients, or any condition deemed by the investigator to make the participant unsuitable for the study were also grounds for exclusion.

### Sample size

A total of 72 participants were screened in the study of which 70 were randomized into 2 groups with 35 participants each. A total of 3 participants dropped out of the study (2 in *C. fistula* extract group and 1 in control group). None of the participants dropped out due to occurrence of adverse events due to consumption of study product or procedure. The details of participant enrollment, allocation, follow-up, and analysis are illustrated in the CONSORT flow diagram.

**Figure 1:**
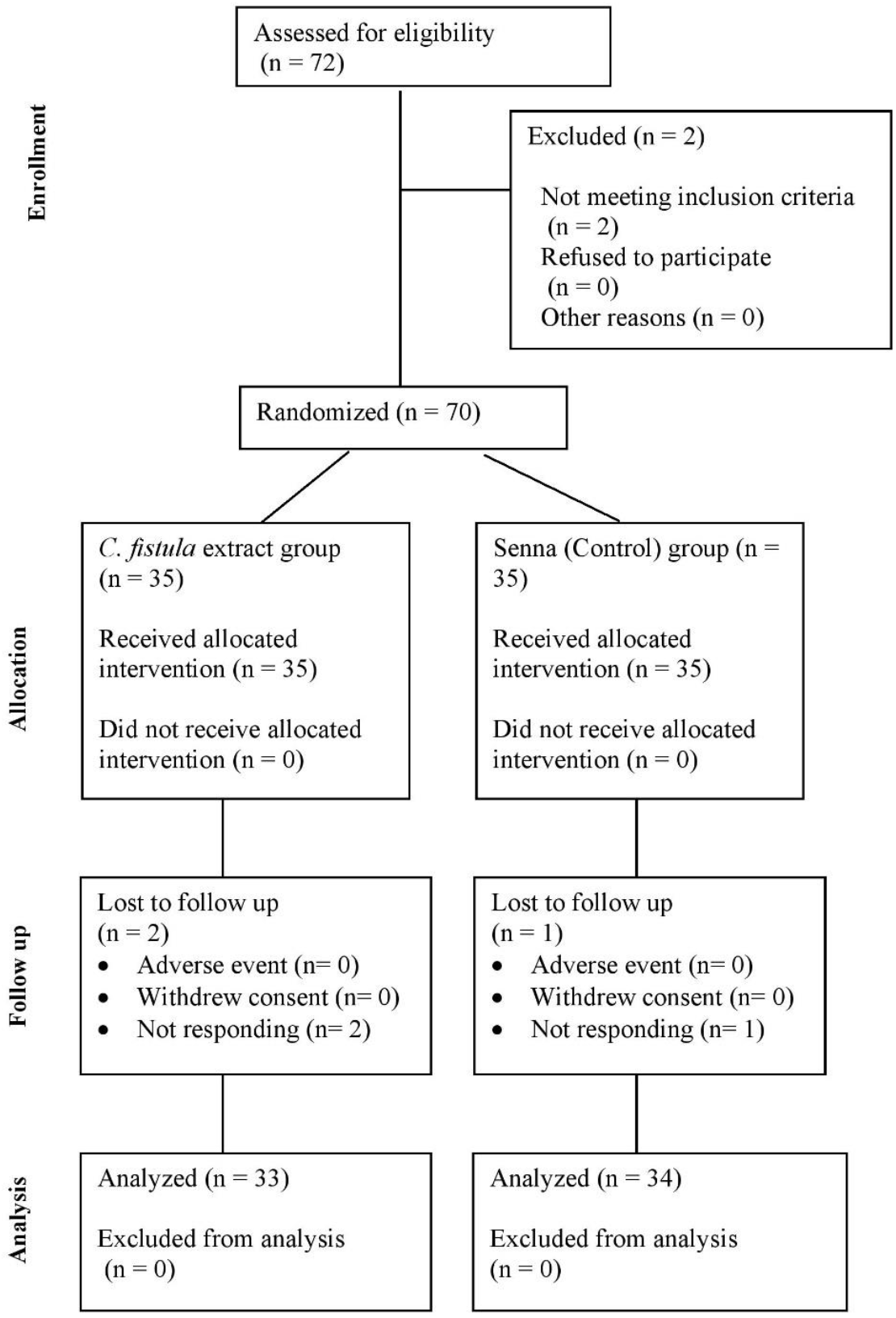
Consort diagram.

### Outcome measures

The primary outcome was comparative assessment of changes in the frequency of bowel movements from baseline to the end of the study and between the two groups. Secondary outcomes included comparative assessments of changes in stool form using the Bristol Stool Form Scale^13^; symptoms of constipation measured using the VAS scale such as straining, bowel satisfaction after defecation, sensation of anorectal blockage, use of manual manoeuvres, average time spent on defecation, and changes in time to first bowel evacuation post-intervention (based on participant diary cards). Additional assessments comprised of associated clinical symptoms including headache, belching, flatulence, abdominal distension, and acidity measured using the VAS scale—all evaluated from baseline to the end of the study, global overall efficacy as rated by both investigators and participants, and product safety and tolerability – evaluated through vitals, laboratory parameters, adverse events and overall safety. Comparative evaluation of these parameters – including CBC, ESR, Hb%, fasting blood sugar, liver function, and renal profile – was conducted at screening and at the end of the intervention.

### Statistical analysis

Statistical analysis was performed Graphpad statistical software. For the analysis of efficacy variables, data was analyzed from the Intent to treat population and per protocol population also. The values of the last visit were considered for final analysis for participants who did not complete the study schedule (Last Observation Carry Forward) for intent to treat analysis. Safety Analysis was done on all participants who have administered at least one dose of treatment. Data describing quantitative measures were expressed as mean ± standard deviation or standard error or the mean with range. Qualitative variables were presented as counts and percentage. Comparison of variables representing categorical data was performed using t test and Chi-square test. All *P* values were reported based on two-sided significance, and all the statistical tests were interpreted at least up to 5% level of significance.

## Results

### Baseline Demography

Both groups were comparable in terms of age, sex, body weight and BMI.

**Table 1:**
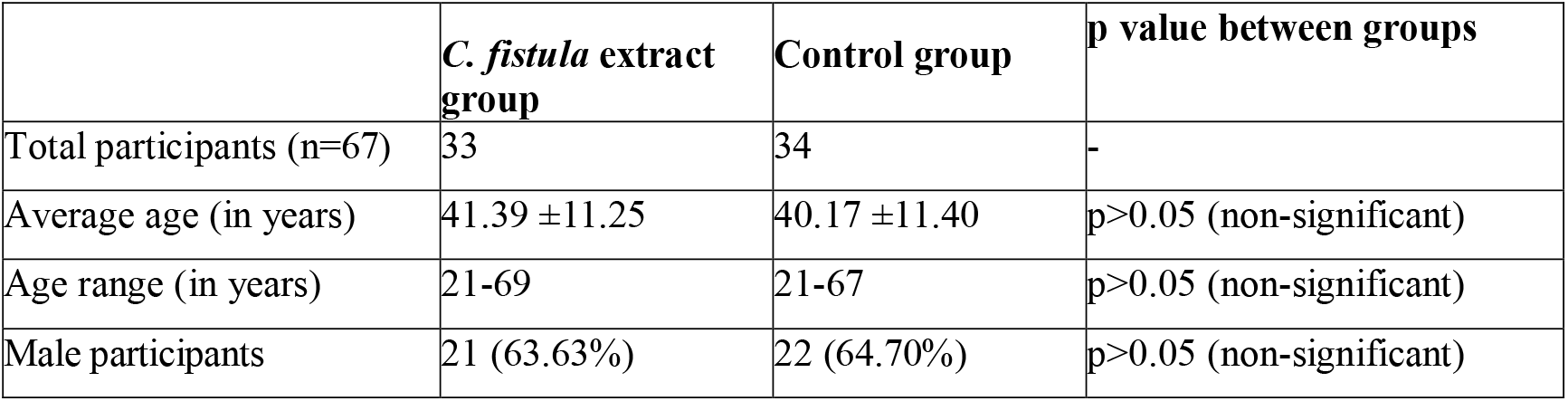
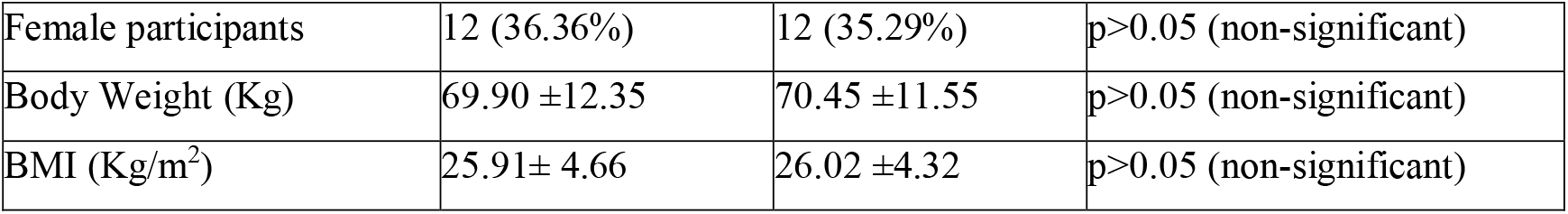
Baseline Demography.

|  | ***C. fistula* extract group** | **Control group** | **p value between groups** |
| --- | --- | --- | --- |
| Total participants (n=67) | 33 | 34 | - |
| Average age (in years) | 41.39 ± 11.25 | 40.17 ± 11.40 | p>0.05 (non-significant) |
| Age range (in years) | 21-69 | 21-67 | p>0.05 (non-significant) |
| Male participants | 21 (63.63%) | 22 (64.70%) | p>0.05 (non-significant) |
| Female participants | 12 (36.36%) | 12 (35.29%) | p>0.05 (non-significant) |
| Body Weight (Kg) | 69.90 ±12.35 | 70.45 ±11.55 | p>0.05 (non-significant) |
| BMI (Kg/m<sup>2</sup>) | 25.91± 4.66 | 26.02 ±4.32 | p>0.05 (non-significant) |

### Frequency of bowel movements

In the *C. fistula* extract group, the mean weekly bowel movement frequency increased significantly from 3.30 ± 0.92 at baseline to 5.90 ± 1.24 on Day 14, with a sustained effect of 4.60 ± 1.19 at Day 21 post-discontinuation. Intergroup analysis showed no significant difference, indicating that the standardized *C. fistula* extract offers comparable efficacy to Senna.

**Table 2:**
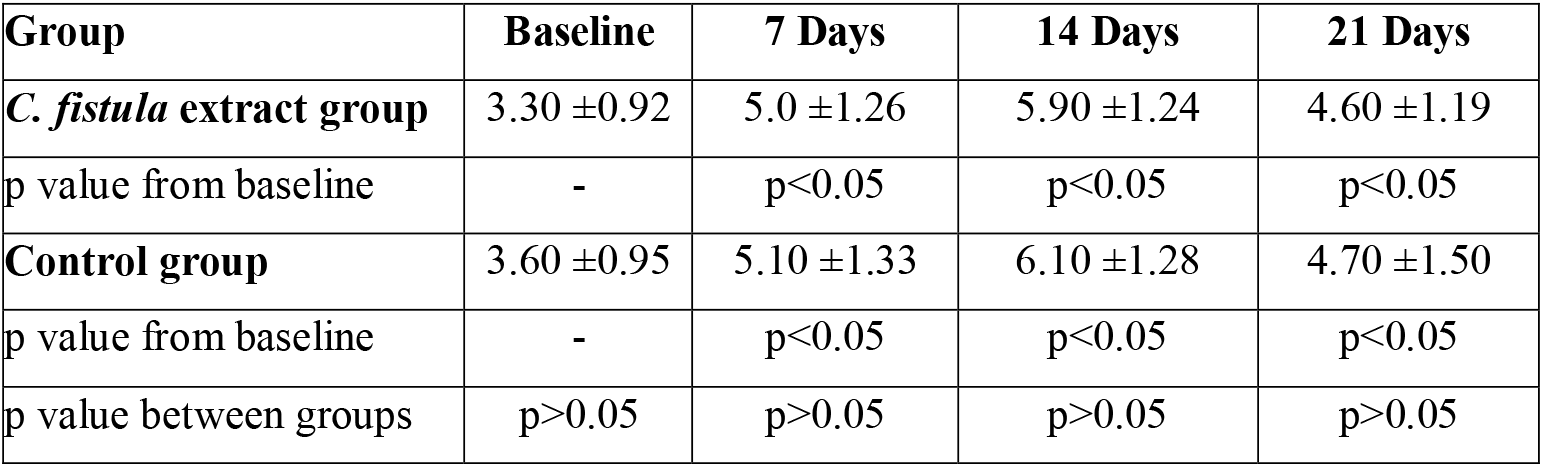
Frequency of bowel movements.

| Group | Baseline | 7 Days | 14 Days | 21 Days |
| --- | --- | --- | --- | --- |
| ***C. fistula* extract group** | 3.30 ±0.92 | 5.0 ±1.26 | 5.90 ±1.24 | 4.60 ±1.19 |
| p value from baseline | - | p<0.05 | p<0.05 | p<0.05 |
| **Control group** | 3.60 ±0.95 | 5.10 ±1.33 | 6.10 ±1.28 | 4.70 ±1.50 |
| p value from baseline | - | p<0.05 | p<0.05 | p<0.05 |
| p value between groups | p>0.05 | p>0.05 | p>0.05 | p>0.05 |

### Bristol Stool Scale based stool consistency

*C. fistula* extract significantly improved Bristol Stool Scale scores over the study period, indicating effective stool softening, which sustained post-discontinuation. These effects were observed to be statistically comparable to Senna.

**Table 3:** Bristol Stool Scale based stool consistency.

| Group | Baseline | 7 Days | 14 Days | 21 Days |
| --- | --- | --- | --- | --- |
| ***C. fistula* extract group** | 2.71 ±0.55 | 3.53 ±0.72 | 3.80 ±0.76 | 3.41 ±0.73 |
| p value from baseline | - | p<0.05 | p<0.05 | p<0.05 |
| **Control group** | 2.75 ±0.61 | 3.58 ±0.81 | 3.87 ±0.81 | 3.45 ±0.85 |
| p value from baseline | - | p<0.05 | p<0.05 | p<0.05 |
| p value between groups | p>0.05 | p>0.05 | p>0.05 | p>0.05 |

### Straining on defecation

Straining on defecation (assessed using the VAS scale) significantly decreased over the study period in the *C. fistula* extract group, with improvements maintained after 7 days post-discontinuation. When compared with Senna, intergroup analysis revealed no statistically significant difference, indicating comparable outcomes.

**Table 4:** Straining on defecation.

| Group | Baseline | 7 Days | 14 Days | 21 Days |
| --- | --- | --- | --- | --- |
| ***C. fistula* extract group** | 54.25 ±16.72 | 40.25 ±18.91 | 28.45 ±21.70 | 41.22 ±19.01 |
| p value from baseline | - | p<0.05 | p<0.05 | p<0.05 |
| **Control group** | 55.10 ±18.55 | 43.25 ±19.19 | 32.90 ±21.56 | 46.55 ±19.29 |
| p value from baseline | - | p<0.05 | p<0.05 | p<0.05 |
| p value between groups | p>0.05 | p>0.05 | p>0.05 | p>0.05 |
| p<0.05 Significant from baseline to follow up and between the groups |  |  |  |  |

### Bowel satisfaction after defecation

Bowel satisfaction scores (assessed using the VAS scale) improved significantly in the *C. fistula* extract group over the study period, with marked reductions maintained at Day 21. The pattern of improvement was observed to be statistically comparable to the control group based on intergroup analysis.

**Table 5:**
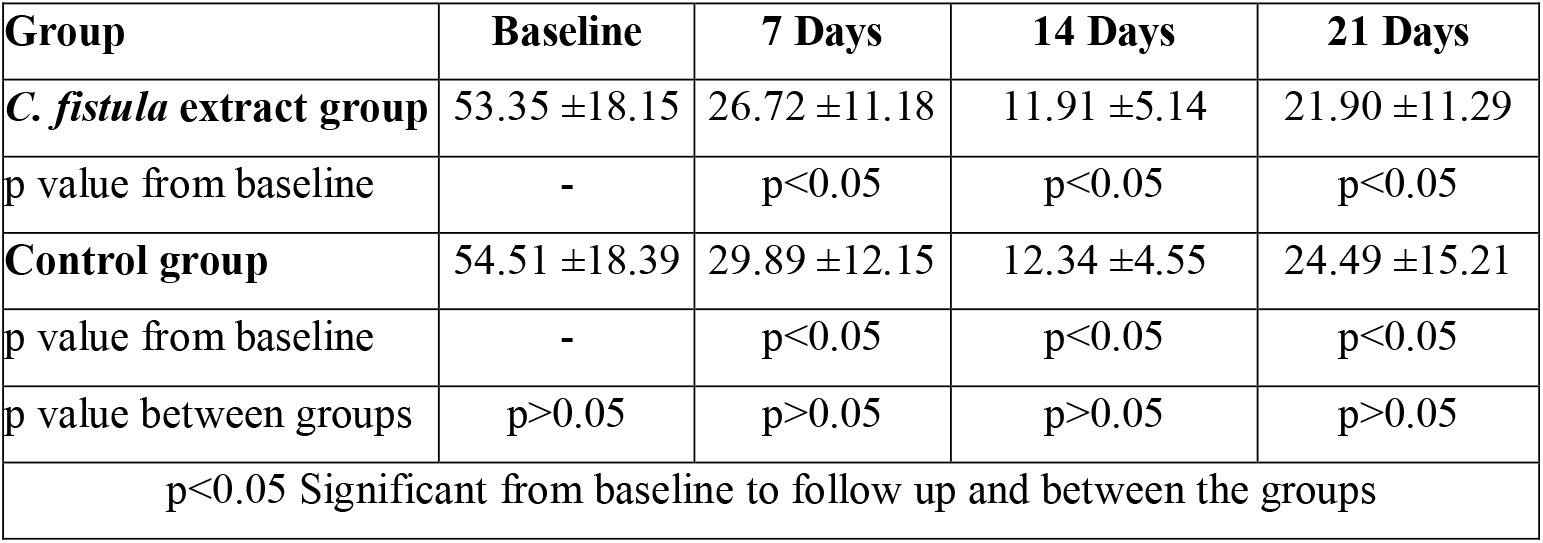
Bowel satisfaction after defecation.

| Group | Baseline | 7 Days | 14 Days | 21 Days |
| --- | --- | --- | --- | --- |
| ***C. fistula* extract group** | 53.35 ±18.15 | 26.72 ±11.18 | 11.91 ±5.14 | 21.90 ±11.29 |
| p value from baseline | - | p<0.05 | p<0.05 | p<0.05 |
| **Control group** | 54.51 ±18.39 | 29.89 ±12.15 | 12.34 ±4.55 | 24.49 ±15.21 |
| p value from baseline | - | p<0.05 | p<0.05 | p<0.05 |
| p value between groups | p>0.05 | p>0.05 | p>0.05 | p>0.05 |
| p<0.05 Significant from baseline to follow up and between the groups |  |  |  |  |

### Sensation of anorectal blockage

Sensation of anorectal blockage (assessed using the VAS scale) decreased significantly in both groups; however, *C. fistula* extract group showed a significantly greater improvement than the control group at Day 14 and and even 7 days post-discontinuation (Day 21), indicating superior efficacy and suggesting its non-habit-forming nature.

**Table 6:** Sensation of anorectal blockage.

| Group | Baseline | 7 Days | 14 Days | 21 Days |
| --- | --- | --- | --- | --- |
| ***C. fistula* extract group** | 39.13 ±12.15 | 22.31 ±12.12 | 15.43 ±9.15 | 19.63 ±11.80 |
| p value from baseline | - | p<0.05 | p<0.05 | p<0.05 |
| **Control group** | 39.18 ±12.81 | 23.67 ±15.19 | 17.83 ±10.89 | 24.48 ±14.91 |
| p value from baseline | - | p<0.05 | p<0.05 | p<0.05 |
| p value between groups | p>0.05 | p>0.05 | p<0.05 | p<0.05 |
| p<0.05 Significant from baseline to follow up and between the groups |  |  |  |  |

### Use of manual manoeuvres

None of the participants in the two study groups required manual manoeuvres during the study period.

### Average time (minutes) spent for defecation

Time spent on defecation decreased significantly in both groups over the study period, with continued improvement observed even after discontinuation. Intergroup analysis showed no statistically significant difference, indicating that *C. fistula* extract is comparable to Senna in reducing the average duration of defecation.

**Table 7:**
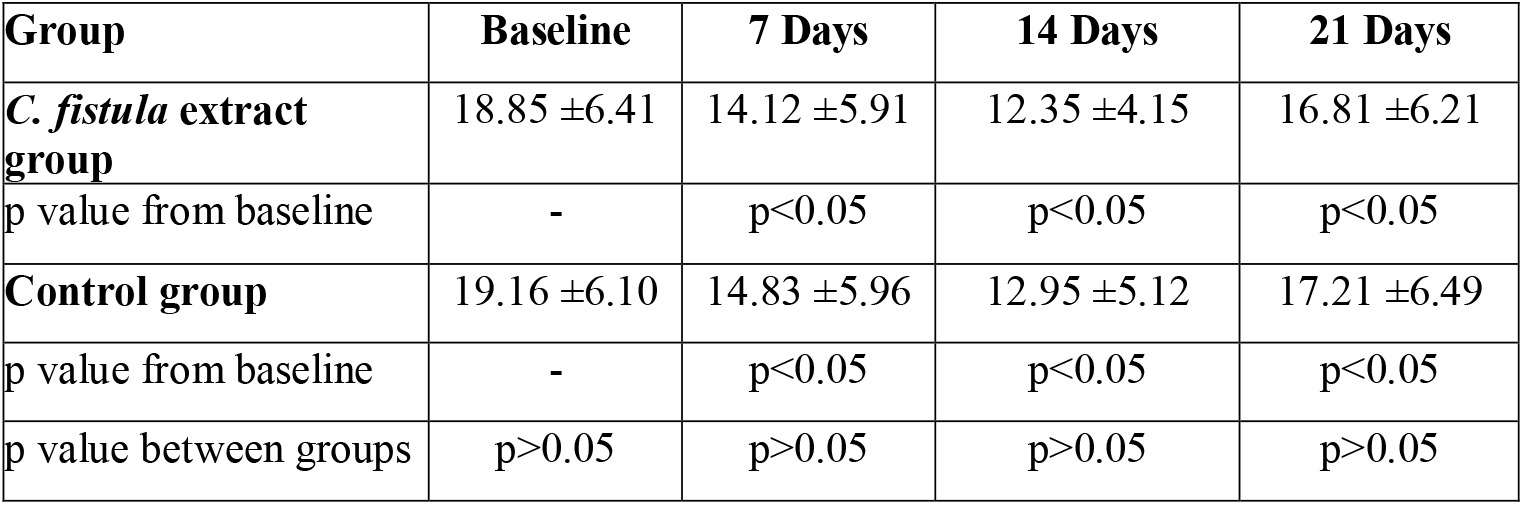
Average time (minutes) spent for defecation.

### Time (hrs) to first bowel evacuation post intervention

*C. fistula* extract group demonstrated a shorter time to first bowel evacuation post-consumption compared to the control group at both Day 7 and Day 14, indicating a faster onset of action.

**Table 8:** Time (hrs) to bowel evacuation post intervention.

| Group | 7 Days | 14 Days |
| --- | --- | --- |
| ***C. fistula* extract group** | 8.75 ±4.00 | 8.12 ±4.10 |
| **Control group** | 9.56 ±4.41 | 9.25 ±4.23 |
| P value between the groups | p<0.05 | p<0.05 |

### Associated clinical symptoms

A significant reduction in symptoms associated with constipation (measured using the VAS scale) viz. headache, acidity, belching, flatulence, abdominal distension, was observed in both the study groups from baseline to 7 days and further to 14 days and 21 days.

### Safety related parameters (vitals, laboratory investigations, overall safety and adverse events)

Both interventions did not affect the vitals and laboratory parameters such as CBC, liver function tests, renal function tests, and urine routine, microscopic examination as demonstrated by statistically comparable values at baseline and day 14 assessments. All parameters were within normal range. *C. fistula* extract was generally rated as having good or excellent overall safety, both by the investigators and the subjects themselves. A total of 8 unrelated adverse events were reported by 7 participants in the *C. fistula* extract group, including cough and cold (n=5), nausea (n=1), and vomiting (n=1). In the control group, 10 unrelated adverse events were reported by 8 participants, comprising common cold with fever (n=4), burning micturition, backache, knee joint pain with swelling, conjunctivitis, dizziness, and burning sensation in the chest (n=1 each). Additionally, 8 participants in the control group reported product-related adverse events such as recurrent abdominal discomfort, abdominal gripping, and sensation of defecation, compared to only 1 participant in the *C. fistula* extract group. All adverse events were mild, self-limiting, and resolved without requiring discontinuation of the study product or any medical intervention.

### Global assessment of overall efficacy as per the physician and participants

While both groups reported high overall improvement, the *C. fistula* extract group demonstrated superior efficacy in participant and investigator assessments. Investigators rated 45.45% of *C. fistula* participants as “very much improved” compared to 38.23% in the control group. Similarly, 48.48% of *C. fistula* participants reported “very much improved,” while only 35.29% in the control group did. Furthermore, no participants in the *C. fistula* extract group reported only “minimally improved,” unlike the control group (5.88% by investigator, 8.82% by participants), indicating a more consistent and impactful positive outcome for *C. fistula*.

|  | *C. fistula* extract group (n=33) |  | Control group (n=34) |  |
| --- | --- | --- | --- | --- |
| Score/Grade Scale | Overall Change by Investigator | Overall Change by participants | Overall Change by Investigator | Overall Change by participants |
| **0 = Not assessed** | 0 | 0 | 0 | 0 |
| **1 = Very much improved** | 15(45.45%) | 16 (48.48%) | 13 (38.23%) | 12 (35.29%) |
| **2 = Much improved** | 18 (54.54%) | 17 (51.51%) | 19 (55.88%) | 19 (55.88%) |
| **3 = Minimally improved** | 0 | 0 | 2 (5.88%) | 3 (8.82%) |
| **4 = No change** | - | - | - | - |
| **5 = Minimally worse** | - | - | - | - |
| **6 = Much worse** | - | - | - | - |
| **7 = Very much worse** | - | - | - | - |

## Discussion

The prevalence of constipation is increasing due to low-fiber diets, sedentary lifestyles, medication use and the ageing population.^14,15^ It can lead to complications such as hemorrhoids, anal fissures, rectal prolapse, etc. and also negatively impact mental health.^16^ These concerns underscore the need for safe and effective treatments, driving interest in alternatives like herbal remedies, which are often seen as having fewer side effects than conventional laxatives ^17,18^

*C. fistula* has been a significant component of traditional medicine, particularly in Ayurveda, for the treatment of constipation. The efficacy of *C. fistula* in managing constipation is primarily attributed to its anthraquinone content, notably sennosides.^19^ These compounds stimulate the colonic smooth muscles, increase peristalsis and facilitate the movement of stools through the lumen, thus promoting defecation. In addition, fluid secretion into the intestinal lumen may be enhanced, contributing to stool softening. ^20^ Beyond these stimulant properties,

*C. fistula* also contains mucilage, a soluble fiber found in various parts of the plant, including the fruit pulp.^21^ Mucilage contributes to its laxative effect by increasing the water content in the intestinal lumen and stool bulk and further stimulating peristalsis to facilitate easier passage.^22^

In this randomized, open-label clinical trial, *C. fistula* extract demonstrated significant improvements across multiple constipation-related parameters over 21 days, with efficacy comparable to Senna. Both groups were demographically comparable at baseline, with no significant differences in age, sex, body weight, or BMI, supporting the internal validity of the comparison. *C. fistula* extract significantly improved bowel movement frequency and stool consistency, with effects sustained even after discontinuation – indicating a potentially longer-lasting benefit. Straining during defecation and bowel satisfaction after defecation were also markedly improved in both groups, and *C. fistula* extract showed a statistically significant advantage over Senna in alleviating the sensation of anorectal blockage by Days 14 and 21. Notably, *C. fistula* extract exhibited a faster onset of action, with a shorter time to first defecation post-consumption compared to Senna. The average time spent on defecation reduced significantly in both groups, with no intergroup difference. None of the participants required manual manoeuvres during the study, and both groups reported a significant reduction in associated clinical symptoms. *C. fistula* extract also demonstrated a favorable safety profile, with no significant changes in vital signs or laboratory parameters. Both groups reported minor, self-limiting adverse events unrelated to the interventions. Some intervention-related discomforts such as abdominal gripping were reported by one participant in the *C. fistula* extract group, in contrast to their observation in as many as eight participants in the control group. The global efficacy assessments further reinforced *C. fistula*’s advantages, showing a higher proportion of “very much improved” ratings and a complete absence of “minimally improved” cases compared to the control group, suggesting a more consistently profound therapeutic effect. Overall, *C. fistula* extract tablets emerged as a safe, effective, and well-tolerated herbal option for the management of functional constipation, offering comparable, and in some areas, superior outcomes to conventional Senna. The sustained efficacy post-discontinuation also suggests that *C. fistula* extract may be a non-habit-forming alternative, warranting further investigation in longer-term studies.

## Conclusion

The study indicates that *C. fistula* extract significantly improved bowel movement frequency, softened stool, and reduced straining during defecation, demonstrating efficacy comparable to Senna. It also proved superior in alleviating the sensation of anorectal blockage and achieving a faster time to first bowel evacuation post-consumption. Its benefits, including improved bowel parameters and reduced straining, were sustained post-discontinuation. Patients also reported significantly reduced time spent on defecation and markedly improved bowel satisfaction. Constipation-associated symptoms—headache, acidity, belching, flatulence, and abdominal distension—were significantly alleviated by the extract comparably to the control group. *C. fistula* also exhibited a more favorable safety profile, with significantly fewer product-related adverse events. and no adverse effects were observed on vitals or laboratory parameters. Therefore *C. fistula* extract can be safely and effectively recommended for the management of constipation and irregular bowel habits.

## Data Availability

All data produced in the present study are available upon reasonable request to the authors

## Notes

### Competing Interest Statement

The authors have declared no competing interest.

### Clinical Trial

CTRI/2024/08/072838

### Funding Statement

This study did not receive any external funding

### Author Declarations

Study was conducted after approval from the Institutional Ethical Committees of the respective study sites- D. Y. Patil University, School of Ayurveda - DYPUSA/24/351 Nirvikar Hospital - ECR/1876/INST/MH/2023 Seth Govindji Raoji Ayurved Mahavidyalaya - IEC/LTR/SGR/12/2024-25 CTRI registration was done with registration number - CTRI/2024/08/072838. Consenting participants meeting the inclusion criteria were considered for the study. The study was carried out and reported adhering to CONSORT Statement.

